# Acute Pain Pathways: protocol for a prospective cohort study

**DOI:** 10.1101/2021.10.21.21265156

**Authors:** Molly Moore Jeffery, Mitra Ahadpour, Summer Allen, Richardae Araojo, Fernanda Bellolio, Nancy Chang, Laura Ciaccio, Lindsay Emanuel, Jonathan Fillmore, Gregg H. Gilbert, Patricia Koussis, Christine Lee, Heather Lipkind, Celeste Mallama, Tamra Meyer, Megan Moncur, Teryl K. Nuckols, Michael A. Pacanowski, David B. Page, Elektra Papadopoulos, Jessica D. Ritchie, Joseph S. Ross, Nilay D. Shah, Mat Soukup, Christopher O. St. Clair, Stephen Tamang, Sam S. Torbati, Douglas W. Wallace, Yueqin Zhao, Rebekah Heckmann

## Abstract

**Introduction:** Opioid analgesics are often used to treat moderate-to-severe acute non-cancer pain; however, there is little high-quality evidence to guide clinician prescribing. An essential element to developing evidence-based guidelines is a better understanding of pain management and pain control among individuals experiencing acute pain for various common diagnoses.

**Methods and analysis:** This multi-center prospective observational study will recruit 1,550 opioid-naïve participants with acute pain seen in diverse clinical settings including primary/urgent care, emergency departments, and dental clinics. Participants will be followed for 6 months with the aid of a patient-centered health data aggregating platform that consolidates data from study questionnaires, electronic health record data on health care services received, prescription fill data from pharmacies, and activity and sleep data from a Fitbit activity tracker. Participants will be enrolled to represent diverse races and ethnicities and pain conditions, as well as geographical diversity. Data analysis will focus on assessing patients’ patterns of pain and opioid analgesic use, along with other pain treatments; associations between patient and condition characteristics and patient-centered outcomes including resolution of pain, satisfaction with care, and long-term use of opioid analgesics; and descriptive analyses of patient management of leftover opioids.

**Ethics and dissemination:** This study has received approval from IRBs at each site. Results will be made available to participants, funders, the research community, and the public.

**Trial registration number:** NCT04509115

**Strengths and limitations of this study:**

- This study addresses a key knowledge gap by recruiting a large and diverse group of patients with acute pain, following them for 6 months, and prospectively assessing their pain experience and acute pain treatment
- Patients will be recruited from multiple treatment settings that are important in the treatment of acute pain, including emergency departments, primary care, dental settings, and surgical settings.
- A patient-centered health data aggregating platform reduces the patient burden of participating in the trial by allowing participants to complete questionnaires when it is convenient and by automatically collecting data on physical activity, healthcare service use, and prescription fills
- All participants will have been offered an opioid prescription, reducing the ability to compare outcomes across opioid and non-opioid pain medications
- Despite a large sample size, there may be limited numbers of specific acute pain diagnoses with a sufficient sample to analyze and compare by condition/diagnosis

## INTRODUCTION

Opioid analgesics are often used to treat moderate-to-severe acute non-cancer pain; however, there is little high-quality evidence to guide clinician prescribing. Many studies report the efficacy of opioids for acute pain, but few address the ideal dose and duration of treatment for various types of pain. Among the many clinical guidelines that recommend a specific duration of treatment or number of pills, most rely on expert consensus in the absence of higher quality evidence.^1^ Appropriate opioid prescribing for acute pain must balance both potential benefits and risks to patients and broader risks to public health.^2^

### Current Knowledge

There is no consensus on the definition of acute pain, but it is broadly considered to be pain that is time limited and of sudden onset.^3^ Acute pain is often treated differently depending on the practice setting in which patients find themselves and the specific providers from whom they receive care. For example, a recent study of post-surgical pain conducted at Mayo Clinic demonstrated substantial variation across surgeons in the amount of opioids prescribed (measured in milligram morphine equivalents [MMEs]) which could not be explained by patient factors for 25 of the most common surgical procedures.^4^ This finding was replicated using national data from insurance claims.^5^ The large observed variability across surgeons in opioid volume prescribed, even after accounting for patient characteristics, suggests that clinicians lack evidence-based guidance on opioid prescribing for post-surgical pain. A recent systematic review found that studies of post-surgery opioid prescribing commonly reported 50% to 70% of prescribed opioid tablets were left unused.^6^ These unconsumed opioids suggest overprescribing and pose a potential risk to patients and the community if the opioids are diverted for non-medical use.

Data are limited with regard to the experience of acute pain, rates of opioid prescribing, clinical outcomes, alignment with recommendations, and disparities in care, particularly outside of surgical care, such as emergency departments (EDs), primary care/urgent care settings, and dental and oral surgery centers.

### Emergency Department

An analysis of the Medical Expenditure Panel Survey from 1996-2012 indicated that overall opioid prescribing increased 471% during that time but that the contribution of prescriptions from EDs decreased from 7.4% to 4.4%.^7^ While more recent data demonstrate a high level of adherence to recommended guidelines for prescribing of opioids in the ED,^8^ there is still limited evidence around the best approaches for pain control and opioid use after initial prescription in the ED. Additionally, the guideline recommendations for dose and duration of treatment are largely based on expert opinion and lower-quality evidence.^9-14^ Furthermore, a recent meta-analysis and systematic review found that Black and Hispanic patients in US EDs were less likely to receive treatment for acute pain than their White counterparts; few studies assessed data for Asian and Native American patients, making the estimates less robust, but the data suggested there may be similar disparities.^15^

### Primary Care/Urgent Care

Although primary care providers may treat a larger proportion of patients with chronic pain than their surgical or emergency medicine colleagues, providers in primary care and urgent care settings still treat a diverse array of conditions associated with acute pain, including headaches, dental pain, and musculoskeletal injuries. Preliminary studies have suggested that prescriptions for 7 or fewer days of opioid medications are adequate for most patients^16^ seen in primary care. However, a study of primary care visits for common acute pain conditions found that 46% of the opioid prescriptions filled after these visits were for more than 7 days’ supply, and 10% were for 30 or more days’ supply.^17^ Additionally, when US primary care providers prescribe opioids, they are the most common type of provider to provide a long-term prescription, with 40% of these initial prescriptions exceeding a 7-day supply.^18^

### Dental Care and Oral Surgery

A recent study comparing opioid prescribing by dentists practicing in the US and UK found that US dentists were much more likely to prescribe opioids than UK dentists (58.2 opioid prescriptions per US dentist per year vs. 1.2 opioid prescriptions per UK dentist per year).^19^ This large difference may indicate that US dentists are overprescribing opioids, but without additional detail on patients’ use and disposal of their dental opioid prescriptions, we cannot determine what harms may be associated with this presumed over-prescription. Despite this comparatively high rate of opioid prescribing by US-based dentists, dentists are less likely to prescribe long courses of opioids when compared with other types of prescribing providers in the United States.^18^

Opioid prescribing to adolescents may be associated with an increased risk of later substance use disorders and overdoses. For example, a population-based study in Sweden found a 30% to 40% relative increase in a composite measure of substance-related morbidity compared to matched cohorts not receiving an opioid.^20^ In the United States, dentists were the most common prescribers of opioids for people aged 0 to 21 years, writing 38.2% of these prescriptions.^21^ Among adolescents and young adults (age 12 to 25), tooth extractions were by far the most common procedure associated with a new dental opioid prescription (79.4%).^22^{Chua, 2021 #1129}

It is not known whether dental prescribing is likely to progress to long-term opioid use or whether unused opioids from dental prescribing are likely to be retained.

### STUDY AIMS

The purpose of this study is to ascertain and describe the trajectories of pain experienced by opioid naïve patients who are prescribed an opioid analgesic for acute pain. Using a digital health-based patient-centered data aggregation platform, we aim to characterize patterns of use of opioids and other pain medications. We aim to enroll a total of 1,550 patients receiving primary and urgent care, emergency department care, or dental care in 5 healthcare systems, along with several community dental practices. Opioid naïve patients offered a new short-acting opioid prescription for acute pain will be recruited and followed prospectively for 6 months to assess pain trajectories, analgesic and non-pharmacologic treatment use, activity, and health care service use. A patient-centered health data aggregating platform (Hugo Health; further information below) will be used to collect patient-reported outcomes and structured data from pharmacy and electronic health record (EHR) patient portals as well as patient-generated data collected through personal activity tracking devices (Fitbit).

There are three study aims:

1. To assess patients’ pain and opioid use patterns in episodes of acute pain for which opioids were prescribed, tracking pain severity and persistence, as well as other prescription and over-the-counter pain medication use
2. To examine associations between patient demographic, clinical and emotional characteristics and outcomes of pain severity and persistence, opioid and non-opioid treatment patterns, satisfaction with care, and barriers to care
3. To assess how patients handled unused opioids both during and at the end of treatment (i.e., leftovers)

## METHODS

This is a prospective cohort study using a novel, patient-centered electronic health data aggregating platform to follow study participants for 180 days, gathering rich data on the course of acute pain, how people treat their pain, and how this acute pain affects their social and emotional functioning.

Opioid naïve patients offered an opioid prescription for acute pain will be recruited at the point of care where the prescription was written. They will be followed for 180 days, during which they will use their personal smartphone or other web-connected device to answer questionnaires that track the location, severity, and daily consequences of their pain; treatments they are using to manage their pain; treatment effectiveness; mood; and potential indicators of problematic opioid use. Structured information from EHR and pharmacy patient portals will be collected to record prescriptions that were written and filled, healthcare service use, and healthcare service-related outcomes (e.g., emergency department visits).

The study will be based at five primary investigative sites in the United States (Figure 1): Yale University (Connecticut), Mayo Clinic (Minnesota), University of Alabama at Birmingham, Monument Health (South Dakota), and Cedars-Sinai Medical Center (Southern California). Participants will be recruited at both community and academic hospitals and clinics associated with these sites, along with community dental practices in several southeastern states along the southern Appalachian Mountains (enrollment coordinated through University of Alabama at Birmingham). Recruitment will stop when approximately 1550 subjects have been enrolled. Enrollment targets (Table 1) are stratified by study site and care setting (emergency department, primary and urgent care, and dental care), with overall demographic targets to ensure the study population is diverse in both racial and ethnic representation and rural/urban residence. Rural dwelling will be determined using participant home ZIP code. Race and ethnicity will be determined by participant self-report, offering the opportunity for participants to select multiple races. People who self-define as part of a racial or ethnic minority and as white will be classified by their non-white race or ethnicity for purposes of counting enrollment quotas.

**Figure 1:**
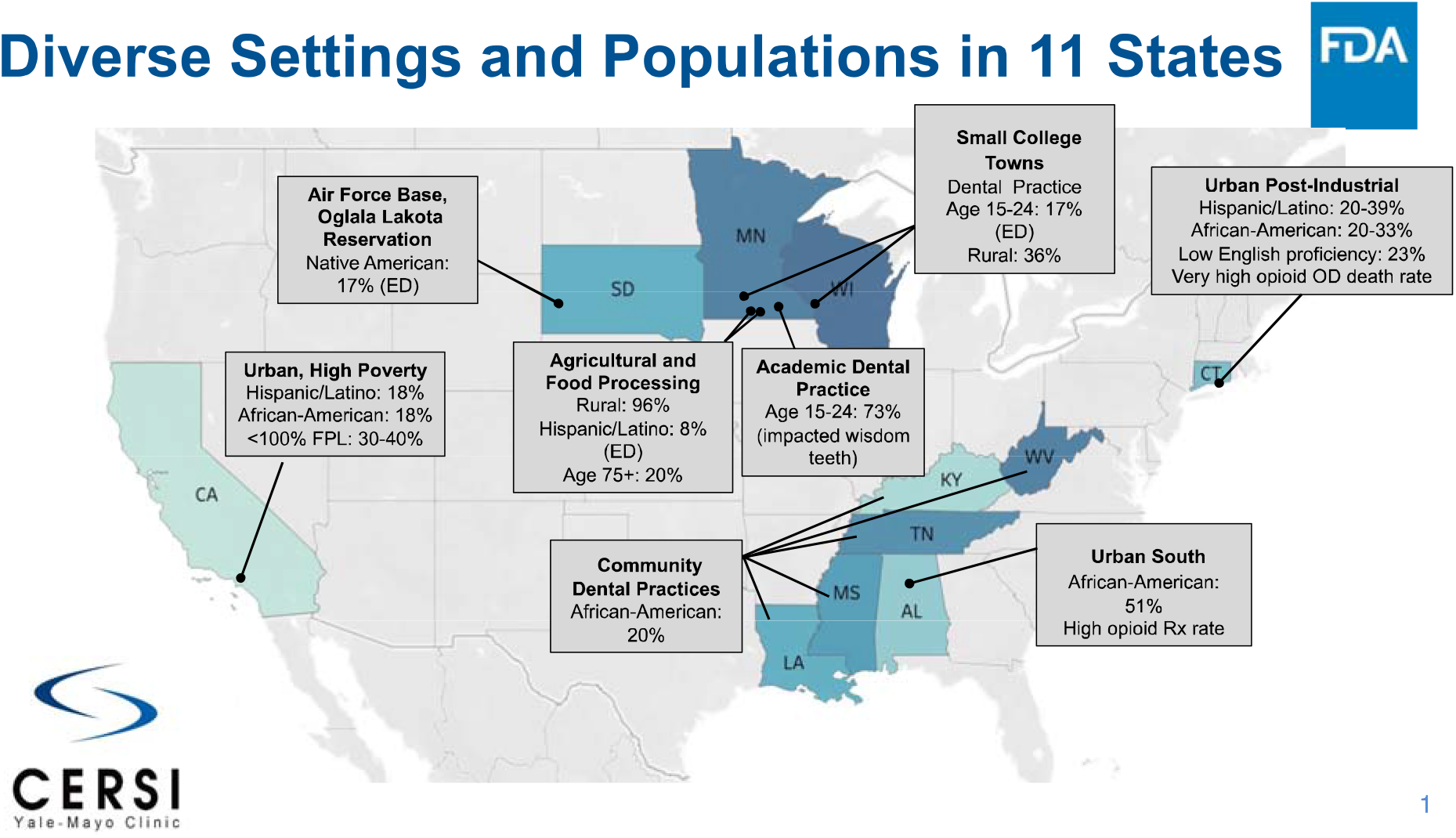
Study sites.

**Table 1:** Planned patient enrollment.

|  | Mayo Clinic | University of Alabama Birmingham | Yale-New Haven Health | Monu ment Health | Cedars-Sinai | Community Dental Practices | Total |
| --- | --- | --- | --- | --- | --- | --- | --- |
| Emergency Department | 100 | 100 | 150 | 150 | 200 | 0 | 700 |
| Primary and Urgent Care | 100 | 100 | 150 | 150 | 0 | 0 | 500 |
| Dental | 100 | 100 | 0 | 0 | 0 | 150 | 350 |
| **Total** | **300** | **300** | **300** | **300** | **200** | **150** | **1550** |
| Overall demographic goals: 20%+ racial or ethnic minorities<br>20%+ rural |  |  |  |  |  |  |  |

**Table 2:**
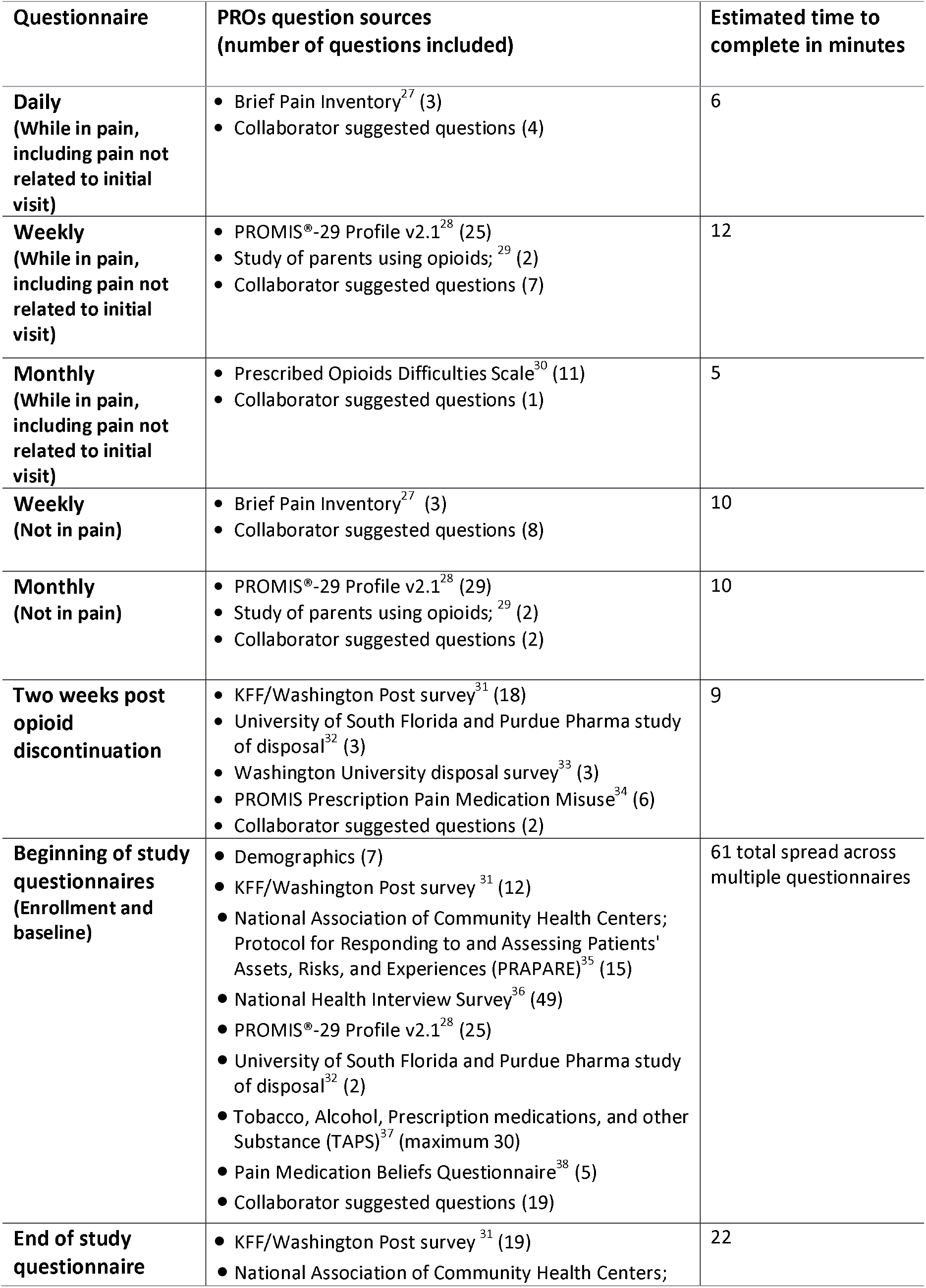
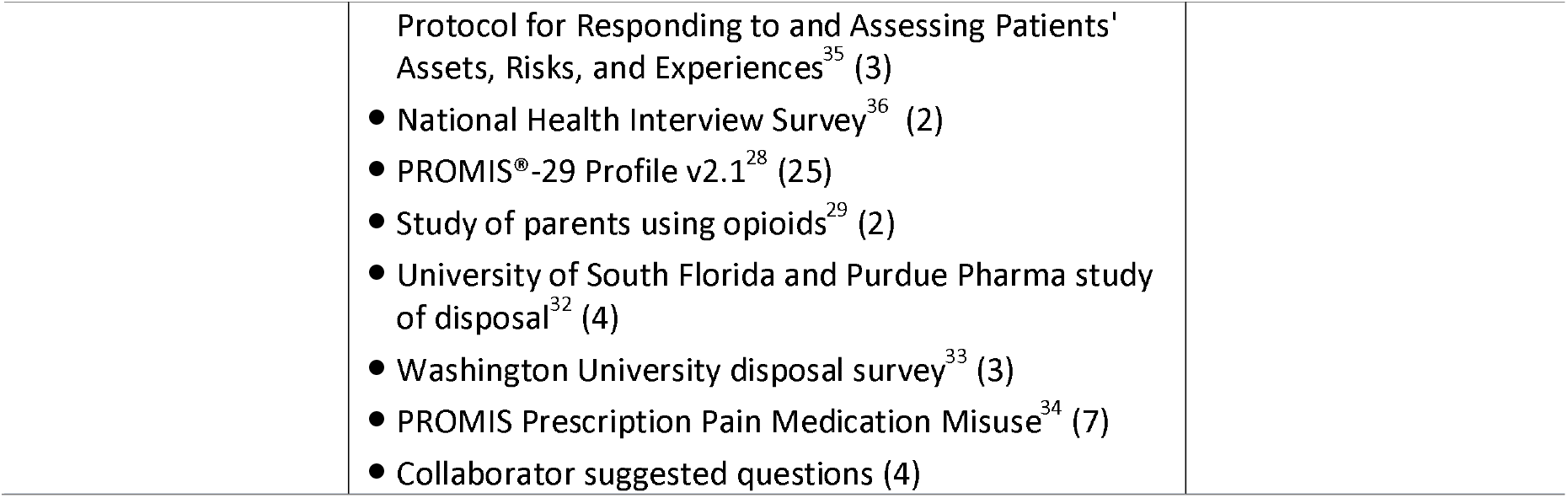
Content of study questionnaires.

**Table 3:** Data extracted from electronic health record, pharmacy portals, and wearable devices.

| Source | Measure/data | Purpose/use |
| --- | --- | --- |
| **Electronic health record** | Current Health Issues | Record of comorbidities for use in understanding risk factors |
|  | Current medications | Additional detail on comorbidities and risk factors (e.g., coprescription of benzodiazepines) |
|  | Appointments and visits | Healthcare service use for related outcomes (e.g., emergency department visits, pain management visits, physical therapy) |
|  | Medications administered | Record of administered medications for treatment of pain (e.g., IV or sub-Q opioids given in ED, steroids, etc.) |
|  | Medications prescribed | Details of pain-related prescriptions (medication, dose, number prescribed) |
| **Pharmacy portal** | Medications dispensed | Details of pain-related prescriptions; determine whether written prescriptions were filled |
|  | Other current medications | Additional detail on comorbidities and risk factors (e.g., coprescription of benzodiazepines, potentially by providers outside the health system) |
| **Fitbit** | Steps/activity | Details on daily movement in relation to patient-reported pain |
|  | Sleep | Details on sleep patterns in relation to patient-reported pain |

### Inclusion Criteria

1. Age 18 and over; or age 15 and over undergoing third molar extraction at Mayo Clinic in Rochester;
2. English-speaking;
3. Experiencing an acute pain condition of less than 8 weeks duration at the time opioids are offered;
4. Offered prescription for opioid analgesic to treat acute pain;
5. Opioid naive (no use of prescribed opioids or illicit opioids, including medical or non-medical use, in the past 6 months) by self-report;
6. Willing and able to give consent and participate in study;
7. Able to access a mobile device (smartphone or tablet) or computer with web access to complete study questionnaires; able to connect Fitbit to a device that can regularly link to Hugo for data transfer;
8. Willing to use the health data aggregating platform;
9. Released/discharged to home after their visit.

### Exclusion criteria

1. Pain thought by treating clinician to be caused by a systemic disease very likely to progress to chronic pain (e.g., sickle cell disease, fibromyalgia, lupus, multiple sclerosis, etc.);
2. Cancer or end-of-life related pain;
3. Unable to give consent and be enrolled within 3 days of being offered the prescription.

It is a limitation of the study that we are not able to provide smartphones or tablets to participants who do not have one. However, smartphone adoption is high at 85% of American adults, including 76% of those with income below $30,000 and 80% of those living in rural areas, and with minimal gaps by race and ethnicity.^23^

## Measures and data collection

### Overview of electronic health data aggregating technology

Hugo is a patient-centered health data aggregating platform developed to foster partnership between research study/clinical trial participants and investigators. Hugo currently enables the secure, automatic, and ongoing deposit of three types of data: clinical data via retrieval from patient portals; patient-generated data with support for a growing number of wearable/medical devices; and patient-reported data via a flexible survey feature. Hugo survey links can be delivered directly to users by email or text enabling the collection of encrypted responses for a wide range of questions including validated patient-reported outcome measures. Proprietary software harmonizes data from different sources, aligning data types and eliminating duplicate information. By leveraging each person’s right to access his or her own data, Hugo’s patient portal-based approach solves patient identity matching issues and removes the need for data sharing agreements with health systems and institutions.

Questionnaires will be sent by Hugo to patients at varying intervals throughout the follow-up period, with the specific questions and frequency dependent on patient-reported pain and opioid use.

- All patients will receive:
  - An enrollment questionnaire to complete during the enrollment process
  - A baseline questionnaire to be completed at home
  - A closeout questionnaire sent at the end of the 6-month follow-up period
- While patients report pain (including pain that is not related to their initial visit), they will receive:
  - Short daily questionnaires
  - Weekly questionnaires
  - Monthly questionnaires
- When patients are not in pain, they will receive:
  - Weekly questionnaires
  - Monthly questionnaires

When patients report in their daily questionnaires that pain is resolved (for 3 days in a row the patient reports an overall pain rating of 0), the daily questionnaires will be stopped, and patients will instead only receive weekly and monthly questionnaires. Two sets of weekly and monthly questionnaires are available; one for those actively reporting pain and one for those not reporting pain. The appropriate questionnaires will be sent to patients based on their response to previous questionnaires. Should they begin reporting pain again in a weekly questionnaire, the “in pain” questionnaire schedule will resume until they again report they have stopped experiencing pain, or until the follow-up period ends.

Two weeks after patients report that they have stopped taking an opioid to treat their pain, they will receive an additional questionnaire that will examine reasons for opioid discontinuation, pain treatment satisfaction, medication disposal, and whether they experienced any potential indicators of medication misuse. Patients continuing opioid use through the end of the follow-up period will be asked the questions on pain treatment satisfaction and medication misuse at the end of the follow-up period.

Using the Hugo platform, we will collect data from electronic medical records from each participating system, all other locations where patients receive care, and from pharmacy portals (Walgreens, CVS, and Walmart) related to medication fills. Together, this will allow us to capture information on comorbidities, opioid and non-opioid analgesics prescribed, and prescription opioids and non-opioid analgesics dispensed.

### Analysis plan

An important goal of this project is to develop evidence to support condition-specific prescribing guidelines that address common acute pain conditions for which opioids are prescribed.

#### Primary analyses

The primary statistical analyses will be descriptive analyses of the outcomes derived from patient questionnaires describing the acute opioid analgesic use of participants. Each outcome will be summarized across care setting populations (i.e., primary care, dental care, emergency department) and also stratified by race and ethnicity, rurality, gender, age group, and opioid indication/patient condition.

##### Specific Aim 1

To assess patients’ pain and opioid use patterns in episodes of acute pain for which opioids were prescribed, tracking pain severity and persistence, as well as other prescription and over-the-counter pain medication use

- Distribution of initial opioid prescription and subsequent dispensings: drug type, strength, number of pills dispensed, and directions for use
- Use of non-opioid drugs/treatments: drug or treatment, number of days used, number of days used on which opioids were also used (i.e., overlap with opioid treatment)
- Comparison of directions for use and actual use: for each day the person takes opioids, measure whether the number of pills taken falls within the range prescribed, below the prescribed range, or above the prescribed range
- Time to opioid discontinuation: defined as 30 days with no opioid use. The time to discontinuation will be counted from initial opioid use to the day on which the last opioid was taken. If 30 days pass with no opioid use, but the participant takes an opioid on the 31st day, a new episode of opioid use will be considered to have started and the time to opioid discontinuation will again be assessed.
- Time to pain resolution (defined as patient no longer indicating pain in the body area initially treated)
- Number of opioid dispensings and total amount of opioids dispensed (in MME)
- Average steps per day registered by Fitbit and trajectory over time (decreasing, increasing, stable) for the periods between enrollment and stopping opioids, enrollment and pain resolution

##### Specific Aim 2

To examine associations between patient demographic, clinical and emotional characteristics and outcomes of pain severity and persistence, opioid and non-opioid treatment patterns, satisfaction with care, and barriers to care

- Reported satisfaction with health care received to treat pain
- Reported barriers to accessing additional treatment: proportion reporting difficulty getting a refill, getting a pharmacy to fill, getting insurance to pay for treatment, being able to afford to pay for treatment
- Progression to long-term use of opioids
- Progression to chronic pain
- Possible misuse of opioids (percent reporting use of opioids for purpose other than pain relief, percent reporting use of opioids for longer than directed or using more than directed, percent using opioids not prescribed for them, etc.); proportion reporting each indicator

##### Specific Aim 3

To assess how patients handled unused opioids both during and at the end of treatment (i.e., leftovers)

- Received health care provider or pharmacist instructions on disposal: proportion reporting receiving information on how to properly dispose of medication, receiving information on the importance of disposing of unused medication
- Opioids left over after opioid discontinuation and estimated amount used during study period (measured in absolute amounts—e.g., mg of oxycodone—as well as MME and tablets)
- Method of storage used
- Method of disposal used
- Motivations for disposing of or keeping leftover drugs (proportion reporting each response [see questionnaire for responses])

#### Secondary analyses

Secondary analyses will assess the association between patient characteristics and outcomes using statistical modeling and will be exploratory in nature.

Patient characteristics that will be included in the analyses:

- Clinical comorbidities, including other chronic and acute painful conditions, as reported by the participant in the beginning of study questionnaire
- Patient age and other demographics
- Substance use
- Pain severity ratings (average from first week of follow-up; time-varying covariate)
- Pain interference
- Treatment effectiveness (i.e., did the patient report that opioids were effective)
- State of residence

### Data Collection and Management

The Hugo platform will collect all information available in the EHRs, pharmacy records, and through the wearable devices and questionnaires. No data will be collected by investigators directly from the health systems at which patients are receiving care. All linked health portals are password-protected and all account links are verified by the study participant upon set up. Patients authenticate themselves through the use of their unique device account username and password.

The data will reside within the Hugo cloud-based platform and will be transferred to Mayo Clinic via a secure file transfer service. The data will then reside on secure servers at Mayo Clinic. The analytic data files stored at Mayo Clinic will not be de-identified but will be treated as a limited data set where patient identifiers are stored in a separate linkable file. The analytic data files will have other PHI such as dates of service, etc. and will be used for enrollment, troubleshooting, and data monitoring.

The data collected as part of this project will not be part of the medical record and will not be provided back to the clinical care team. Patients will be made aware of this as part of the informed consent process and will be asked to contact the care team directly if they have any concerns related to their health.

Data will be stored at Mayo Clinic for five years after the end of the study. At the end of the five years, data will be archived in secure storage at Mayo Clinic similar to clinical trial and other prospectively collected data.

De-identified data will be stored at the FDA indefinitely for sharing both with internal investigators as well as external researchers. The informed consent will be explicit in that participants are agreeing for their de-identified data to be used by external groups for research or regulatory purposes.

#### Confidentiality

All data and records generated during this study will be kept confidential in accordance with Institutional policies and HIPAA on subject privacy.

#### Data Quality Monitoring

The research staff will regularly monitor the status of the portal data (EHR/pharmacy/Fitbit) coming into the Hugo platform using the version of the Hugo dashboard only available to research staff. Within this dashboard, the research staff will be able to review the connection status of all of the portals connected to each patient’s Hugo account. Should a connectivity issue be noticed by research staff, they will note the connection issue reported by the dashboard and determine if the issue is limited to one participant or if multiple participants are experiencing the same problem. The research staff will then follow up directly with the Hugo support team. Once the source of the issue is identified, the research staff will follow-up with the affected patients as needed to correct the issue in a timely fashion.

Research staff will also keep track of any technical issues directly reported by patients during their follow-up period. If the technical issues are not able to be resolved by the research staff, or if multiple patients report the same problem, the research staff will forward the issue to the Hugo support team to identify and correct the issue and in order to follow-up with those patients affected.

The study was selected to receive quality monitoring oversight using the FDA Office of Translational Sciences Quality Management System (QMS) for human subject research (HSR) conducted or supported by the FDA. The QMS involves the use of standard operating procedures (SOPs), checklists and templates when conducting quality assurance monitoring activities including site visits. In 2020, 4 virtual pre-enrollment visits (due to COVID-19 restrictions) were completed as well as 1 onsite interim visit in 2021. Individuals contracted to monitor the study on behalf of the FDA may access personal information collected as part of this study, including all of the information shared with the research team from the patients’ Hugo account. Monitoring activities are done to ensure adequate projection of the rights, welfare, and safety of human subjects and the quality of the clinical trial data. These individuals are all obligated to maintain confidentiality by the nature of their work or are bound by confidentiality agreements.

### Statistical Methods

#### Baseline Data

Baseline and demographic characteristics will be summarized by standard descriptive summaries (e.g., means and standard deviations for continuous variables such as age and percentages for categorical variables such as race and ethnicity, medians and quartiles for skewed data).

#### Primary analyses

The primary analyses will include all subjects. Tables will be presented with percentages, means and standard errors, medians and 25^th^, 75^th^, and 90^th^ percentiles, and minimums and maximums, as appropriate.

#### Secondary analyses

Secondary analyses will be exploratory and use statistical modeling to estimate the association between patient characteristics and outcomes. Survival analysis techniques will be used to analyze time to event outcomes. Negative binomial regression will be used to analyze count outcomes (opioids leftover, number of refills).

#### Missing data

Multiple imputation will be used to handle missing outcome and covariate data in these analyses. Chained equations will be used to impute each variable with missing data^24 25^. The number of imputations used will be 20 or more for each analysis. Sensitivity analyses will be conducted using missing categories for covariates and including only people with non-missing outcome information.

#### Sample size and statistical significance

This study is designed to gather information rather than to detect statistically significant differences between populations. For that reason, we did not provide a sample size analysis.

## Protocol and registration

This study has been registered at ClinicalTrials.gov (NCT04509115).

## Ethics and dissemination

Each site will obtain local IRB approval. Study results will be disseminated through publications in general and specialty medical journals and conferences.

## STUDY UPDATE

As of this writing, all sites have obtained local IRB approval and are enrolling participants. COVID-19 has severely impacted study enrollment at nearly all sites. Several sites were not allowed to recruit in person for some period of the pandemic. The effect of the pandemic and associated changes in healthcare service use on the results of the study are not yet known and will be an important aspect in analysis of the study results.

Based on priority conditions identified by the National Academies of Sciences, Engineering, and Medicine acute pain prescribing working group,^26^ two study sites were given permission to recruit limited numbers of people receiving hip or knee replacement surgery and people who recently gave birth.

## Data Availability

The data are not available to the public

